# Infrared spectroscopy enables rapid, robust, portable COVID-19 saliva screening based on pathophysiological response to SARS-CoV-2

**DOI:** 10.1101/2021.12.22.21268265

**Authors:** Seth T. Kazmer, Gunter Hartel, Harley Robinson, Renee S. Richards, Kexin Yan, Sebastiaan J. Van Hal, Raymond Chan, Andrew Hind, David Bradley, Fabian Zieschang, Daniel J. Rawle, Thuy T. Le, David W. Reid, Andreas Suhrbier, Michelle M Hill

**Author notes:** These authors contributed equally to this work (Gunter Hartel, Harley Robinson).

## Abstract

Fourier-transform infrared (FTIR) spectroscopy provides a (bio)chemical snapshot of the sample, and was recently proposed for COVID-19 saliva screening in proof-of-concept cohort studies. As a step towards translation of this technology, we conducted controlled validation experiments in multiple biological systems. SARS-CoV-2 or UV-inactivated SARS-CoV-2 were used to infect Vero E6 cells *in vitro*, and K18-hACE2 mice *in vivo*. Potentially infectious culture supernatant or mouse oral lavage samples were treated with ethanol or Trizol to 75% (v/v) for attenuated total reflectance (ATR)-FTIR spectroscopy, or RT-PCR, respectively. The control condition, UV-inactivated SARS-CoV-2 elicited strong biochemical changes in culture supernatant/oral lavage despite lack of replication determined by RT-PCR or cell culture infectious dose 50%. Crucially, we show that active SARS-CoV-2 infection induced additional FTIR signals over the UV-inactivated SARS-CoV-2 infection, which correspond to innate immune response, aggregated proteins, and RNA. For human patient cohort prediction, we achieved high sensitivity of 93.48% on leave-on-out cross validation (n=104 participants) for predicting COVID-19 positivity using a partial least squares discriminant analysis model, in agreement with recent studies. However, COVID-19 patients negative on follow-up (RT-PCR on day of saliva sampling) were poorly predicted in this model. Importantly, COVID-19 vaccination did not lead to mis-classification of COVID-19 negatives. Meta-analysis revealed SARS-CoV-2 induced increase in Amide II band in all arms of this study and recent studies, indicative of altered β-sheet structures in secreted proteins. In conclusion, ATR-FTIR is a robust, simple, portable method for COVID-19 saliva screening based on detection of pathophysiological responses to SARS-CoV-2.

## Introduction

More than two years into the coronavirus disease 2019 (COVID-19) pandemic, there remains a need for globally affordable rapid, field-deployable screening tests to detect active Severe Acute Respiratory Syndrome Coronavirus 2 (SARS-CoV-2) infection. While the current gold standard reverse transcriptase quantitative polymerase chain reaction (RT-qPCR) test is highly sensitive in detecting SARS-CoV-2 RNA, the technical requirements, time to result and the accumulated testing costs are prohibitive in developing countries and for disadvantaged communities. Equipment-free rapid antigen tests with immobilized anti-SARS-CoV-2 antibodies in lateral flow devices have been developed to generate results in 5-20 minutes, but the reported variable sensitivity of such diagnostics remain unaddressed (1, 2), and there is also the imperative need to re-evaluate antibody sensitivity as each new variant emerges.

As an alternative to antibody-based rapid testing, Fourier transform infrared (FTIR) spectroscopy was recently reported as a promising, point-of-care technology for COVID-19 detection using pharyngeal swab or saliva (3-5). FTIR provides a biochemical snapshot of the sample by measuring the vibration of chemical bonds (6). FTIR spectra collected from saliva of COVID-19 patients and healthy controls were used to develop prediction algorithms that demonstrated high predictive accuracy in cross-validation of the same cohort (4, 5) or in an independent cohort (3). FTIR sampling using either transflection (slide mount), or attenuated total reflectance (ATR, directly deposited on highly reflective crystal) was able to distinguish healthy controls from confirmed COVID-19 cases with high specificity and sensitivity (3-5).

These recent cross-sectional cohort studies provided promising proof-of-concept for the use of FTIR in COVID-19 screening using saliva as a non-invasive sample that can be self-collected. To further develop this technology towards point-of-care application, we generated comprehensive data on the pathobiological basis underpinning the SARS-CoV-2/COVID-19 FTIR signal using a rapid and biosafe processing method. We utilized three different biological systems: culture supernatant from *in vitro* cell infection, oral lavage of inoculated hACE-2 mice, and human saliva from a limited cohort of COVID-19 patients and health controls. For the cell and mouse models, UV-inactivated SARS-CoV-2 virus was used as control, and two post-infection time points were examined. The potentially infectious biological samples were first decontaminated by adding 100% ethanol (v/v) to 75% final (v/v), as per our recent study using ATR-FTIR of plasma samples for prediction of COVID-19 disease severity (7). The high ethanol percentage facilitated the rapid evaporation of the treated plasma (1 μl) on the ATR-FTIR target (∼30 sec), which enables very rapid data acquisition (7). In the current study, we used the same procedure to analysed cell culture secretome, mouse oral lavage and human saliva samples, and additionally conducted proteomic analysis of the mouse oral lavage samples to elucidate the pathobiology. Finally, a meta-analysis was conducted of all available COVID-19 FTIR spectra data.

## Results

### Characterisation of *in vitro* SARS-CoV-2 infection-induced secretome ATR-FTIR spectra

As a first step, a standard Vero cell *in vitro* infection model was used to investigate the secretory host response to SARS-CoV-2 infection. Two controls were used: a media control and ultraviolet light (UV)-inactivated SARS-CoV-2, which cannot replicate as UV destroys RNA. RT-qPCR of SARS-CoV-2 RNA of the culture supernatant confirmed the lack of infectivity for both controls while the active infection demonstrated an increased SARS-CoV-2 RNA load at 24 and 48 hrs (Fig 1a).

**Fig 1.**
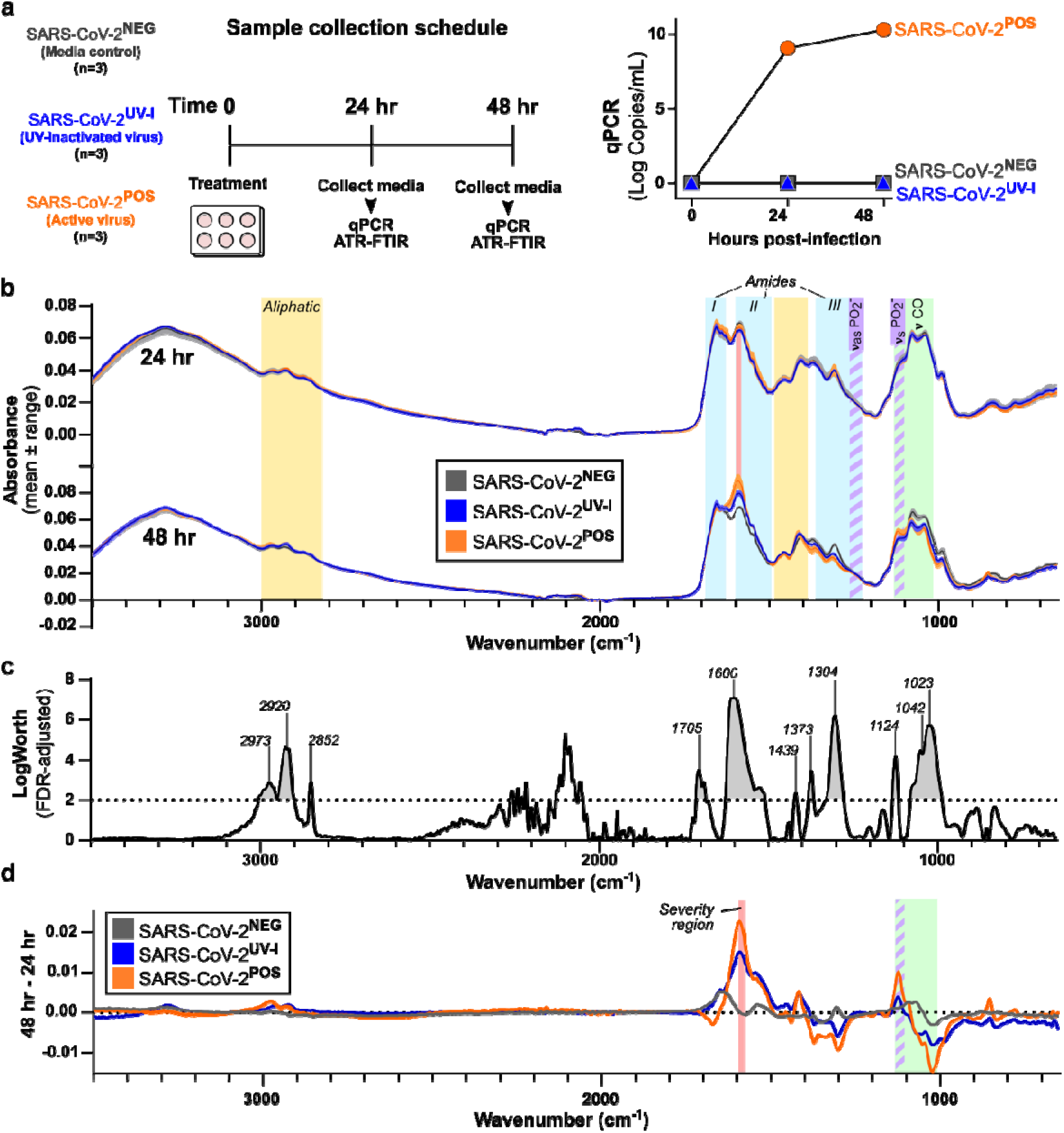
ATR-FTIR spectral changes of culture supernatants, *in vitro* SARS-CoV-2 infection model. **a**) Vero-E6 cells (6×10^5^) were treated with media alone (SARS-CoV-2^NEG^), UV-inactivated (SARS-CoV-2 ^UV-I^), or SARS-CoV-2 (SARS-CoV-2^POS^) for 2 hours, after which cells were washed in PBS and media replaced. Aliquots of conditioned media were collected at 24 hr and 48 hr post-infection for qPCR and ATR-FTIR. Verification of viral load was accomplished via RT-qPCR (p=0.0035). **b**) Overlapping spectra of technical replicates for 24 hr and 48 hr timepoints. Colored bands indicate chemical components of interest: Aliphatic (yellow), Amide I/II/III (cyan), severity(*10*) (red), Saccharide (green), phosphodiester asymmetric stretching (*v*_as_PO_2_^-^) and symmetric stretching (*v*_s_PO_2_^-^) (purple stripes). **c**) Significant features of SARS-CoV-2 infection compared to the two controls at 48 hr, using FDR-LogWorth analysis; dotted line represents FDR-LogWorth 2 (p<0.01). **d**) Subtraction of supernatant spectra per each treatment from 24 hr to 48 hr.

Interestingly, despite the 9 log increase in SARS-CoV-2 RNA at 24 hr, the FTIR spectra of the supernatant showed minimal change at this time point, apart from increased absorbance at Amide I band (1700-1600 cm^-1^) in the active SARS-CoV-2 infected sample (Fig 1b). At 48 hr, the FTIR profiles of UV-inactivated SARS-CoV-2 and active SARS-CoV-2 infected supernatants showed increased bands at 2970 cm^-1^, 2924 cm^-1^, 2874 cm^-1^, 1590 cm^-1^, 1415 cm^-1^, and decreased at 1373 cm^-1^, 1309 cm^-1^, 1042 cm^-1^, 988 cm^-1^ compared to media control (Fig 1b and S1a Fig). However, at 48 hr active SARS-CoV-2 infected secretome displayed separation from both controls in Amide I/II (1700-1470 cm^-1^) and fingerprint (FP) region (1450-600 cm^-1^) (Fig 1b), as well as right shifting to a lower wavenumber at 1668 cm^-1^ to 1595 cm^-1^ (Fig 1d and S1b Fig).

An FDR LogWorth analysis confirmed significance in a number of these wavelengths from both controls, shown as regions above the dotted line in Fig. 1c (p<0.001). To clarify the spectral changes per each condition over time, averaged spectra of the 48 hr time point were subtracted from those at 24 hr (Fig 1d and Fig S1). The greatest separations of spectra between active SARS-CoV-2 and UV-inactivated SARS-CoV-2 occurred at 2977 cm^-1^, 2920 cm^-1^, 1668-1665 cm^-1^, 1595 cm^-1^, 1418 cm^-1^, 1298 cm^-1^, 1122 cm^-1^, 1021 cm^-1^, 854 cm^-1^ (Fig 1d and S1 Fig). Active SARS-CoV-2 infection demonstrated separation from media control at 1600 cm^-1^, 1304 cm^-1^, 1124 cm^-1^, 1042 cm^-1^, and 1023 cm^-1^ (Fig 1c,d). These features notably included increased absorbance at 1124 cm^-1^, a region considered to reflect symmetric stretching of phosphodiester linkages of RNA (8, 9)(*v*_s_PO_2_^-^).

### ATR-FTIR spectra of oral lavage from respiratory SARS-CoV-2-infected mouse model

Next, transgenic ACE2 (K18-hACE2) mice were used to evaluate the ATR-FTIR spectra of oral secretome following respiratory SARS-CoV-2 infection, again comparing to UV-inactivated SARS-CoV-2. Past studies determined K18-hACE2 mice develop lung infection and a respiratory disease resembling severe COVID-19 (10, 11), and have been widely used to evaluate interventions against SARS-CoV-2 infection and disease (12-17).

Oral lavage was collected from anaesthetized mice prior to infection (day 0), and then on days 2 and 4 post inoculation (Fig 2a). Body mass of active SARS-CoV-2 infected mice started declining on day 3, reaching minus 10-15% on day 4 (Fig 2b). Comparison of the average oral lavage ATR-FTIR spectra showed more significant changes on day 4 compared to day 2 (S2 Fig). On day 2, aliphatic, fatty acids 1738 cm^-1^, and (*v*_as_PO_2_^-^) bands showed increased absorbance with mild drop in saccharides between groups (S2a, S2b Fig), however, on day 4, saccharides further dropped with pronounced increases in the amide bands (Fig 2c, 2d and S2a, S2b Fig). Subtraction of spectra on day 4 from day 0 (baseline) allowed visualization of respective time-course changes for the UV-inactivated SARS-CoV-2 infection group (SARS-CoV-2^UV-I^) (Fig 2d, top) and the active SARS-CoV-2 infection (SARS-CoV-2^POS^) (Fig 2d, bottom); additional subtractive analysis was performed between these two groups to observe the unique changes attributed to active SARS-CoV-2 infection (ΔTreatment, Fig 2d, middle). A broad, rising Amide II peak at 1542 cm^-1^ was the most prominent feature resulting from SARS-CoV-2 infection. Although all mice displayed an increase in Amide peaks between days 2 and 4, those with active infection presented a significant Amide II peak and Amide I shift (Fig 2c, 2d and S3 Fig, p = 0.00001).

**Fig 2.**
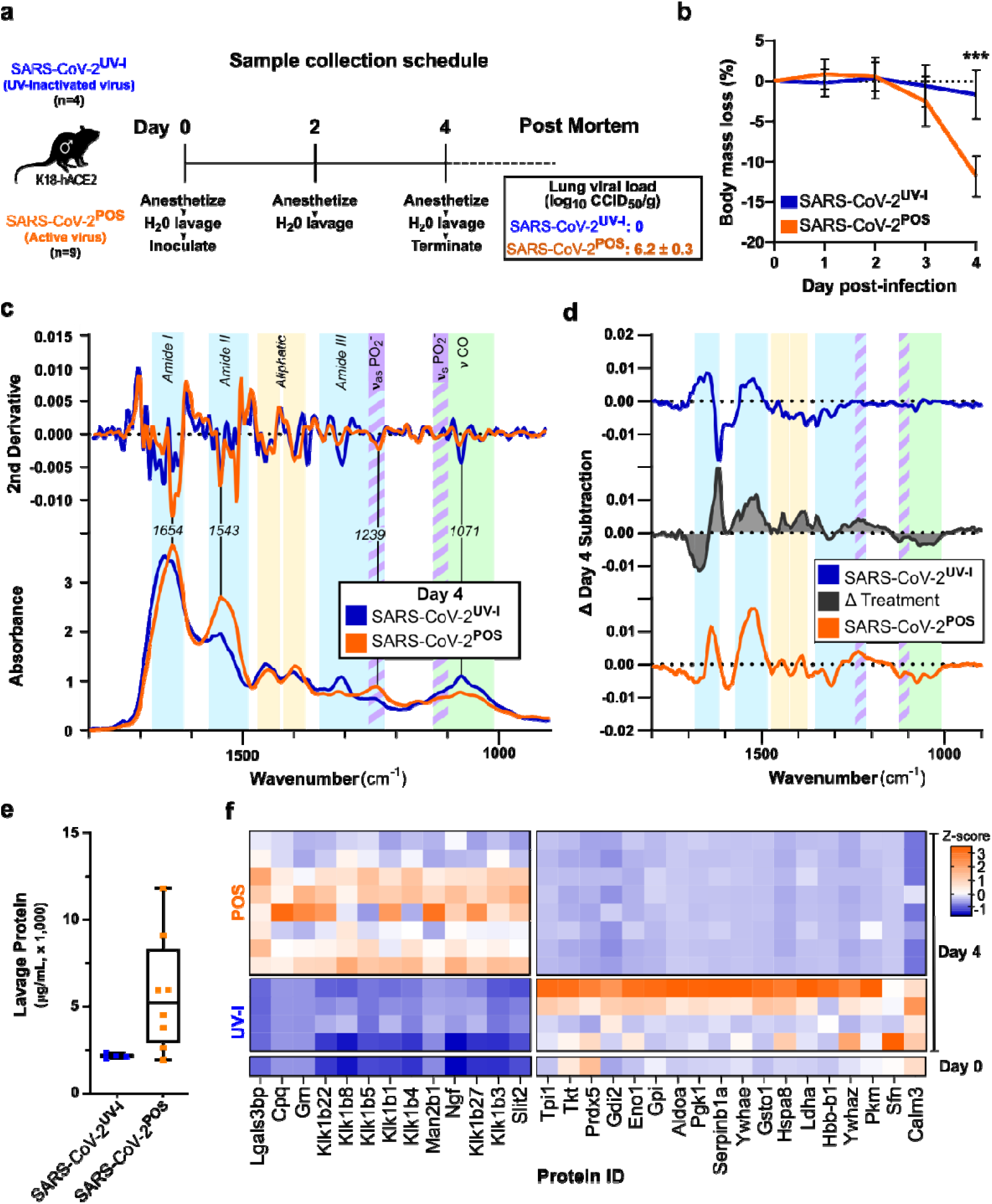
ATR-FTIR spectra and proteomic changes of oral lavage of SARS-CoV-2 mouse model. **a**) Male K18-hACE2 mice were inoculated with intrapulmonary UV-inactivated (n=5) or active SARS-CoV-2 (n=9), and oral lavage was sampled on days 2 and 4. Viral load of mouse lung tissue was assessed via cell culture infectious dose 50% assay (CCID50) post-mortem, showing no active virus in the inactivated virus group. **b**) Body weight measurements were recorded daily. Error bars show standard error. *** on day 4, p= 0.0004. **c**) Day 4 oral lavage ATR-FTIR spectra of the Amide I/II and fingerprint regions with respective 2^nd^ derivative (above). Colored bands indicate chemical components of interest: Amide (protein) bands I, II, III (cyan), PO_2_^-^ asymmetric (*v*_as_) and symmetric (*v*_s_) stretching (purple stripes), saccharides (green), with identification of key peaks by wavenumber. **d**) Subtraction of Day 4 spectra from Day 0, showing a time-course alteration for SARS-CoV-2^UV-I^ (blue) and SARS-CoV-2^POS^ (orange), as well as the difference between the groups, ΔTreatment (black). Complete spectra (4000-600 cm^-1^) as well as Day 2 data are available in Fig. S2. **e**) Protein concentration of Day 4 oral lavage plotted per group. **f**) Proteomics was conducted on equal amounts of Day 4 oral lavage, and a pooled sample of Day 0 oral lavage (3 samples) for comparison. Heatmap shows z-scores of differential proteins (p<0.1 adjusted) between SARS-CoV-2^UV-I^ and SARS-CoV-2^POS^ groups.

To further elucidate the pathophysiology, we conducted untargeted proteomics on the lavage. The protein concentrations of the SARS-CoV-2^POS^ lavage was elevated in comparison to the SARS-CoV-2^UV-I^ group, indicating strong secretory response to SARS-CoV-2 infection (Fig 2e). Proteomic analysis on equal amount of lavage protein revealed upregulation of several kallikreins, and proteins involved in immune modulation such as lectin galactoside-binding soluble 3 binding protein (Lgals3bp) and progranulin (Grn) (Fig 2f). Furthermore, a number of proteins were comparatively down regulated, notably including Calmodulin-3 (Calm3).

### ATR-FTIR spectra of human saliva distinguishes SARS-CoV-2 infection status

To investigate the application of ATR-FTIR for COVID-19 screening in human saliva samples, we collected saliva from 104 participants, healthy controls (COVID.NEG, n=44) and COVID-19 cases (COVID.POS n=60) (Fig 3a, S1 Table).

**Figure 3.**
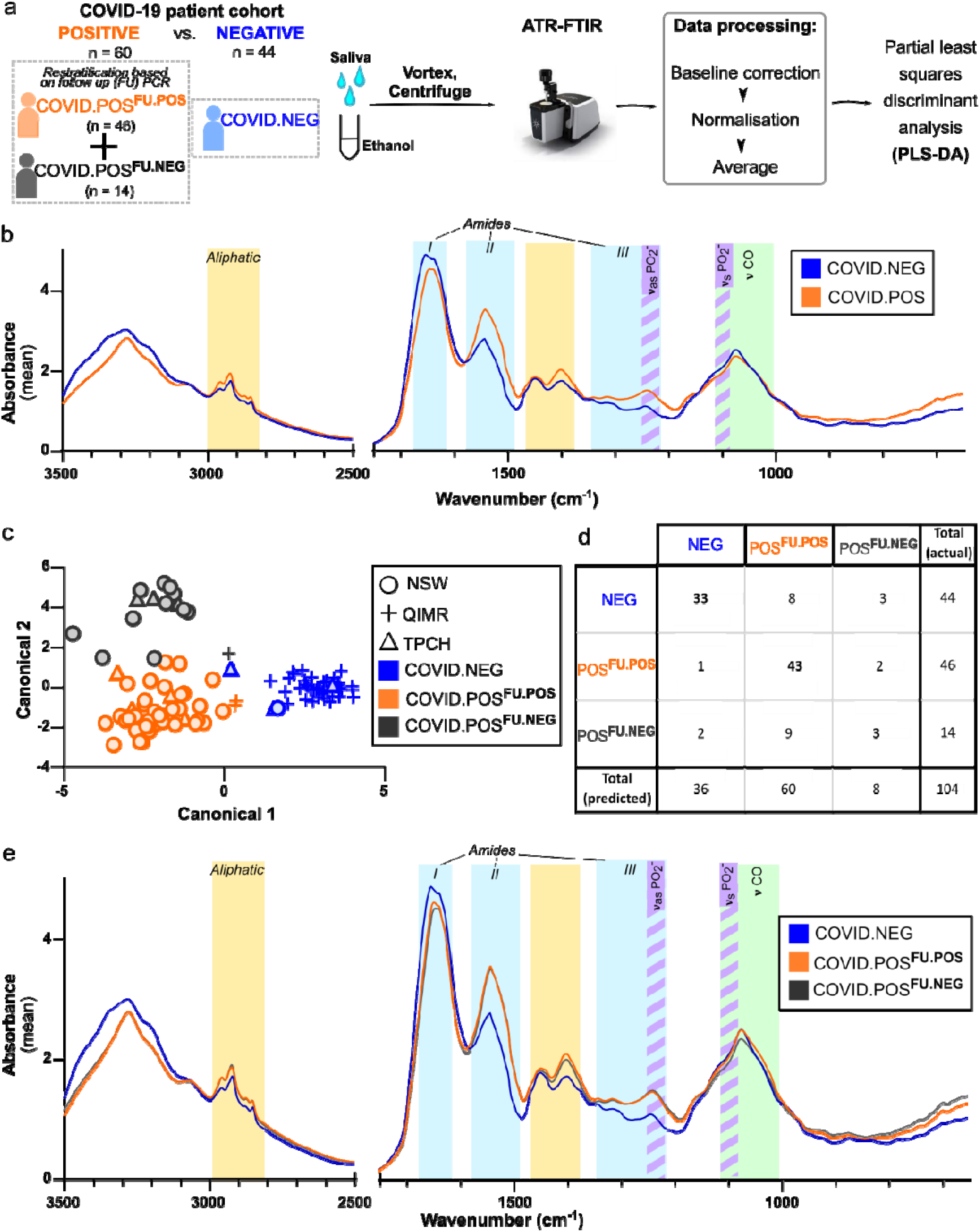
ATR-FTIR spectral data of human cohort. **a**) Workflow. Sublingual saliva were collected from human subjects with known COVID.POS and COVID.NEG status. Follow-up SARS-CoV-2 PCR was conducted for the COVID.POS group on the saliva or swab collected on day of saliva collection (COVID.POS^FU.POS^ or COVID.POS^FU.NEG^). Clarified saliva adjusted to a final concentration of 75% ethanol was used for ATR-FTIR on an Agilent Cary 630, with samples dried (∼30 sec) on the crystal. Data for each technical replicate were baseline corrected then normalized to an AUC of 1. **b**) Average spectra (3500-650 cm^-1^) of COVID.NEG and COVID.POS groups. Colored bands indicate chemical components of interest: Aliphatic (yellow), Amide I/II/III (cyan), Saccharide (green), phosphodiester (purple stripes). **c**) Canonical plot with symbols indicating the location of sampling, NSW, New South Wales Health Pathology; TPCH, The Prince Charles Hospital; QIMRB, QIMR Berghofer Medical Research Institute. **d**) Contingency table for leave-one-out cross-validation of the partial least squares discriminant analysis (PLS-DA) model. Columns represent actual designation while rows represent predicted categorization. **e**) Average spectra (3500-650 cm^-1^) of the three clinical groups.

The acquired spectra (n=3-6 technical replicates per biological sample) were baseline corrected and normalized, then the technical variance in the dataset was assessed using pairwise Euclidean distancing (S4 Fig). The variance within replicates of a participant was significantly lower compared to variance between participants (0.1925 ± 0.1941 vs 0.6089 ± 0.544, p < 0.0001, S4 Fig), indicating acceptable technical variability relative to the observed biological variability.

The average spectra for COVID.POS and COVID.NEG groups showed visible differences in aliphatic, amide I, II, III regions (Fig 3b). Discriminant analysis with canonical plot revealed separation between COVID.NEG and COVID.POS groups on Canonical 1 (X-axis) but interestingly showed separation within the COVID.POS groups along Canonical 2 (Y-axis) correlating to PCR results on the day of saliva sampling; termed COVID.POS^FU.POS^ and COVID.POS^FU.NEG^ (Fig 3c).

A Partial Least Squares Discriminant Analysis (PLS-DA) model using Non-linear Iterative Partial Least Squares (NIPALS) algorithm was developed to predict the three clinical groups. Seven factors explained 75.25% of the variation in the spectra. Based on Leave-One-Out Cross-validation (LOO-CV) the PLS-DA correctly predicted 75% of COVID.NEG (specificity), and 93.48% of COVID.POS^FU.POS^ (sensitivity). In the COVID.NEG group, while the sensitivity was in line with the previous cohort studies, the specificity was slightly lower in this cohort.

Therefore, we investigated if recency of COVID-19 vaccination may contribute to incorrect prediction by saliva FTIR. Of the 44 COVID.NEG participants, 29 participants had received one or both vaccine doses in the 8 -120 days prior to saliva collection. All 4 incorrectly predicted samples had vaccination doses 22-67 days prior to saliva collection suggesting that vaccination did not influence ATR-FTIR results.

Of the 14 COVID.POS^FU.NEG^ participants, only 2 (14.3%) were predicted correctly with the remaining 10 (71.4%) and 2 (14.3%) predicted as COVID.NEG or COVID.POS (Fig 3d). This is not surprising as viral clearance dynamics are highly variable in individual post infection onset.

Although visualisation of the average spectra revealed little separation between these subgroups, a region we previously reported to correlate with COVID-19 disease severity (7) also was able to distinguish between COVID.POS^FU.POS^ and COVID.POS^FU.NEG^ groups (Fig 3e and S5b Fig).

### Delineation of spectral signature for COVID.POS^FU.POS^ saliva

To determine the significant spectral regions between groups, and the regions selected in the PLS-DA model, we conducted Logworth FDR analysis (Fig 4a, 4b) and Variable Importance Plot analysis (Fig 4c), respectively. Compared to COVID.NEG, COVID.POS^FU.POS^ saliva demonstrated significant differences in all amide bands identified: increased absorbance in Amide A and B (3500-3300 cm^-1^, 3100 cm^-1^, respectively), a narrowing of Amide I from a major right shift (1710-1650 cm^-1^) and minor left shift (1624-1596 cm^-1^), a pronounced increase and right shift of Amide II (1570-1470 cm^-1^), and increase of Amide III (1320 cm^-1^). Significantly increased absorbances were also observed in aliphatic bands: 2956 cm^-1^ (*v*_as_ CH_3_), 2870 cm^-1^ (*v*_s_ CH_3_), 1464 cm^-1^ (δ_as_ CH_3_, asymmetric bending), 1420 cm^-1^ (δ CH_2_ and deformations), and 890 cm^-1^ (δ CH_2_). Bordering Amide III, the two most significant combined points, 1252 cm^-1^ and 1228 cm^-1^, represent asymmetric phosphate stretching (*v*_as_PO_2_^-^) among a diversity of macromolecules such as phospholipids, phosphorylated proteins, and RNA (18).

**Fig 4.**
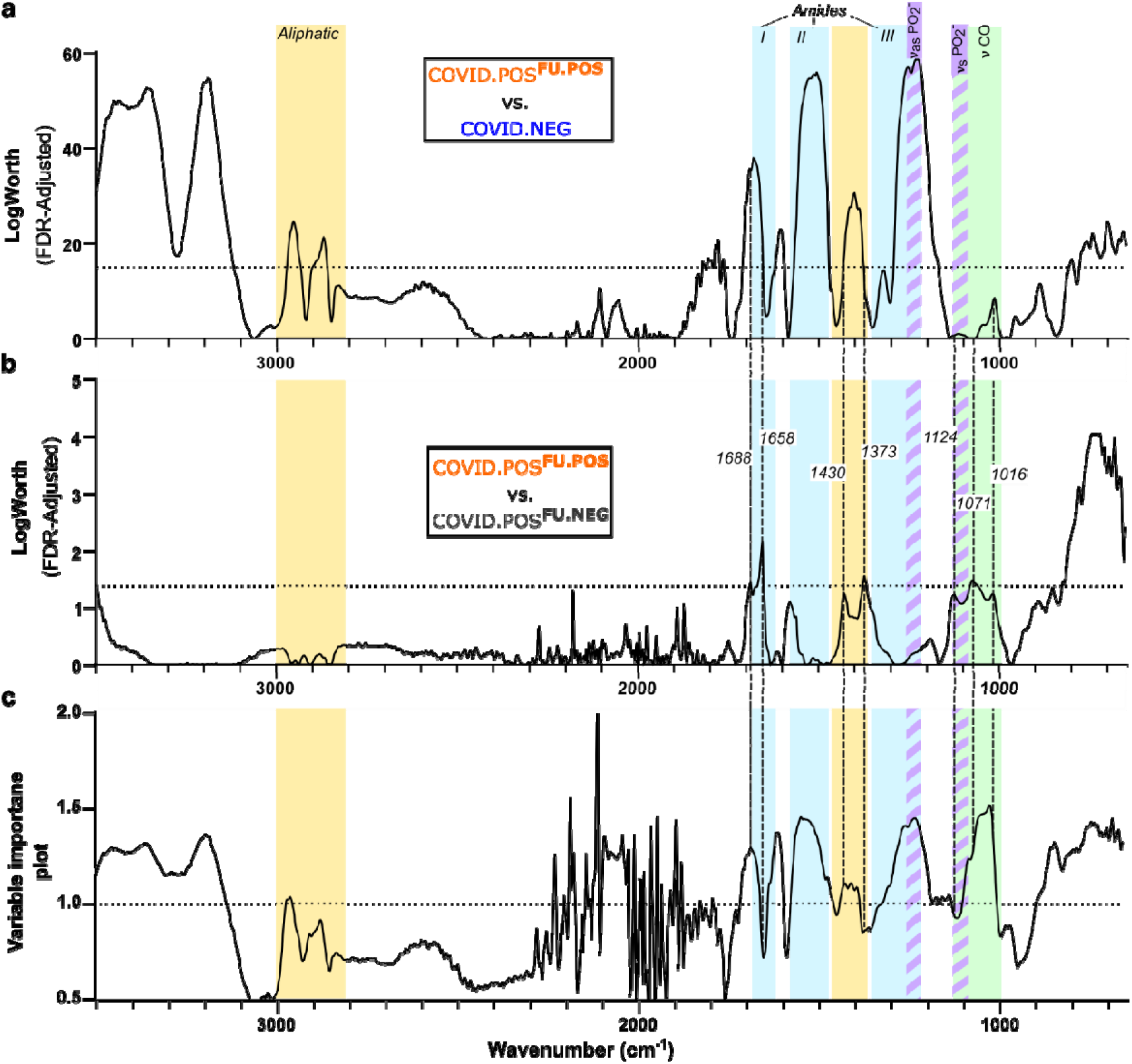
Delineating FTIR spectral signature for COVID-19 saliva screening. Saliva FTIR spectra from the cohort analysis in Fig. 3 was subjected to comparative FDR LogWorth analysis for **a)** COVID.POS^FU.POS^ and COVID.NEG saliva samples, and **b)** COVID.POS^FU.POS^ and COVID.POS^FU.NEG^ samples. Dotted lines visualizes the cut-off chosen at a level above noise for each comparison at LogWorth of 15 (p = 1×10^−15^) and 1.3 (p = 0.05), respectively. **c)** Variable importance plot for the final PLS-DA model shown in Fig. 3d, displaying spectral regions’ contributions to the model. Dotted line indicates VIP of 1.0. Colored bands indicate chemical components of interest: Aliphatic (yellow), Amide I/II/III (cyan), Saccharide (green), phosphodiester (purple stripes).

In addition, 7 points of varying significance (p < 0.06-0.01) correlated with the significant peaks from COVID.POS^FU.POS^ vs COVID.NEG comparison: right shift of Amide I (1688/1658 cm^-1^), decreased aliphatic/RNA (1430 cm^-1^), decreased δCH_3_ - bending (1373 cm^-1^), decreased *v*_s_PO_2_ - RNA (1124 cm^-1^), *v*_s_PO_2_^−^, symmetric and C-O ν ribose (1071 cm^-1^), and decreased *v* C_4_-OH - glucose (1016 cm^-1^). The right shifting Amide I peak in COVID.POS^FU.POS^ compared to both COVID.NEG and COVID.POS^FU.NEG^ is in agreement with residual misfolded amyloid protein fibrils and elevated IgA in COVID-19 patient saliva (S5 Fig)(4, 19, 20).

Comparison of the Logworth FDR analysis with the VIP analysis of the PLS-DA model (Fig 4c) revealed that most of the predictive peaks overlap with the significant peaks from the COVID.POS^FU.POS^ vs COVID.NEG analysis (Fig 4b).

### Delineation of COVID-19 spectral signature across diverse models

Finally, we sought to establish the most characteristic COVID^POS^ spectral signature across multiple models and studies by comparing the significant results from all three study arms, as well as the three recent publications (Table 1), bearing in mind the differing nature of the models and methodologies across studies. This analysis revealed several consistent spectral changes due to SARS-CoV-2 infection across multiple models/studies (Table 1). Most strikingly, a change in the structure of proteins was indicated by Amide II increase in all studies, indicative of β-sheet structures. In all human cohorts (but not *in vitro* or mouse models), Amide III, aliphatic, phosphodiester asymmetric stretching (*v*_as_PO_2_^-^) and saccharide bands were also increased. In contrast, saccharide bands were decreased in *in vitro* and mouse models, with VIP values 1.2-1.43 over the range 1067-1006 cm^-1^ (Fig. 3b/e, Fig. S5b). This range of wavenumbers was previously recognized for having significance in predicting severe COVID-19 outcomes when evaluating blood plasma, including an elevated AUC at 1592-1588 cm^-^1(7). Taken together, these results suggest a change in the physiological response between end-point/severe COVID-19 (*in vitro* cell, mouse model and severity study) and COVID-19 patients who are tolerating the disease well.

**Table 1.** Spectral features for saliva/secretion in COVID^POS^ cohorts/models.

| Band Designation | In Vitro<sup>a</sup> | Mouse<sup>a</sup> | Human<sup>a</sup> | Chemical components (19, 21-26) | Barauna (3) | Wood(5) | Martinez □<br>Quaziti<sup>c</sup>(4) |
| --- | --- | --- | --- | --- | --- | --- | --- |
| Amide A |  | **3246** | 3358<br>3518-3280<sup>b</sup> | N-H, O-H stretching |  | **x** |  |
| Amide B |  | **3067** | 3190<br>3248-3110<sup>b</sup> | Amide II overtone, aromatic amino acids |  | **x** |  |
| Aliphatic | **2973** | 2931-2880<br>2858<br>2837 | **2954**<br>**2870**<br>2968-2944<sup>b</sup> | -CH<sub>3</sub>/-CH<sub>2</sub>; C-H symmetric (v<sub>s</sub>) & asymmetric (v<sub>as</sub>) stretching |  | **x** |  |
| Fatty Acids | 1705 | 1702 | 1722-1704<br>1714-1690<sup>b</sup> | -COOH, C=O v; and ketones |  | **x** | O |
| * Amide I | 1690 |  | 1680<br>1680<sup>b</sup> | Protein β-sheets; C=O guanine |  |  |  |
|  |  | **1638** |  | Protein β-sheets |  |  |  |
|  | **1600** |  | 1625-1594<br>1632-1585<sup>b</sup> | Protein aggregates; amyloid fibrils |  |  | **x** |
| Amide II | **1524** | **1578-31** | **1572-1470** | N-H Mostly β-sheet |  | **x** | **x** |
| Aliphatic Fingerprint |  | 1468 | **1464** | -CH<sub>2</sub> δ, bending vibrations |  | **x** |  |
|  | **1439** | 1431 | **1416**<br>**1420<sup>b</sup>** | -CH<sub>2</sub> δ, symmetric stretching band of carboxyl group, CH<sub>2</sub> ω, wagging; RNA | **x** | **x** | **x** |
|  |  |  | **1402**<br>**1400<sup>b</sup>** | C-H deformation; CH<sub>2</sub> ω; C-N stretching; In-plane C2'OH in RNA |  | **x** | **x** |
|  | 1373 | 1370 | **1375**<br>**1388-1376<sup>b</sup>** | -CH<sub>3</sub> δ, C-H v; methyl bending/stretching |  | **x** | **x** |
| Amide III | 1302 | 1302 | **1319** | Amino acid side-chains; terminal oxygen (PO<sub>3</sub>) |  | **x** | **x** |
|  |  | 1378-1354<br>1335-1280 | **1340-1285**<br>**1330-1177<sup>b</sup>** | -CH<sub>2</sub> ω; -CH<sub>3</sub> δ, amyloid contribution |  | **x** | **x** |
|  |  |  | **1250**<br>**1250<sup>b</sup>** | PO<sub>2</sub> v<sub>as</sub>; C-N v |  | **x** | **x** |
|  |  |  | **1243-1218**<br>**1226<sup>b</sup>** | PO<sub>2</sub> v<sub>as</sub>; amyloid fibrils |  | **x** | **x** |
| RNA | **1124** |  | 1129<sup>b</sup> | PO<sub>2</sub> v<sub>s</sub>, phosphodiester stretching |  | **x** |  |
| Saccharide |  | 1094 |  | -C-O-C, ether linkages; -O-Ca<sup>2+</sup> c | **x** |  |  |
|  | 1072 | 1064 | 1077<sup>b</sup> | PO<sub>2</sub>- v<sub>s</sub>, symmetric and C-O v | **x** | **x** | **x** |
|  | 1050 |  |  | -C-O v, C-OH group; C-C v (sugars) |  | **x** | **x** |
|  | 1023 |  | **1034-1003**<br>**1095-997<sup>b</sup>** | C-O v; P-O v; C-OH δ |  |  | **x** |
|  | 1008 | 1012 | **1012** | C<sub>4</sub>-OH, Glucose |  | **x** | **x** |
|  | 988 | **988-974** |  | PO<sub>2</sub><sup>-</sup> v<sub>s</sub>; -C-O-, ribose |  | **x** |  |
|  |  |  | **940** | P-O v, phosphorylation; -C-C- v |  | **x** |  |
|  |  | **887-866** | **889** | P-O-P v<sub>as</sub>; -C-C- v; aromatics |  | O | **x** |
|  |  | 830 | **836<sup>b</sup>** | P-O-C v; =C-H δ; aromatics |  | O | **x** |

<sup>a</sup> Significance by FDR LogWorth analysis
<sup>b</sup> Also significant by Variable Importance Plot analysis
<sup>c</sup> Long Fasting collection; analysis of truncated spectra
\* Some significance found in Amide I region were due to shift, not proportion change of specific chemical species alone
**Bold type** indicates where SARS-CoV-2 spectra was ↑ higher
X = Separation of spectra from control
O = Noticeable change in spectra, value not legible/not reported

## Discussion

This study provides controlled validation and the underpinning pathobiology data required to support the translation of ATR-FTIR for COVID-19 saliva screening. The consistency of SARS-CoV-2^POS^ FTIR signature across *in vitro*, mouse and human arms of this study, and three existing independent reports (3-5) provides confidence that this robust technology is ready for clinical development and deployment. Compared to the single output of current PCR and antigen tests, the broad biochemical information from the saliva FTIR spectra provides both diagnostic and prognostic information supporting the use of saliva as a non-invasive, self-collectable bio-sample reflective of the physiological response (27-29).

The COVID-19 saliva FTIR signature shares many biochemical features with amyloid deposits (aliphatic, amide, and phosphodiester *v*_as_) and lipofuscin (19, 30). Enhanced amyloid formation in SARS-CoV-2 has been a recent area of focused research (31-33). The consistent COVID-19-associated amide absorbance shifting from α-helix (∼1652 cm^-1^) composition to β-sheet (∼1636 cm^-1^) is likely related to SARS-CoV-2 induction of protein aggregates through its spike protein (34-36). Strikingly, our proteomics data revealed extensive upregulation of kallikrein proteins in oral lavage of SARS-CoV-2^POS^ mice. Savitt et al. recently reported direct interaction and activation of the kallikrein/kinin system (KKS) by recombinant SARS-CoV-2 proteins S, M. N, and E (37). High molecular weight kininogen (HK) and plasma prekallikrein (PK) bring about the sequelae of bradykinin, and complexing of HK/PK with Blood Coagulation Factor XII (FXII) initiate the intrinsic clotting cascade with the aid of misfolded proteins and polyphosphate (38). Polyphosphate may serve as a natural defense blocking the receptor binding domain for SARS-CoV-2 (39). Whether derived from platelets or commensal bacteria, extensive utilization of polyphosphate in the KKS/FXII pathogenesis of SARS-CoV-2 offers another substantial explanation for the phosphate and Amide III profiles among these studies (40-42). Another upregulated protein contributing to the Amide I/II bands, Lgals3bp, is potentially upregulated as a compensatory defensive mechanism to the prolonged innate immune response by day 4 (43). Further investigations should be carried out to establish protein disaggregated in human saliva samples and the involvement of KKS-FXII-polyphosphate.

While the cell culture and mouse model data were generally consistent with our human cohort data, the reduced saccharide band in cell and mouse SARS-CoV-2 treated samples contrasted with the increases observed in human COVID.POS samples. This discrepancy was most probably due to different stages in the evolution of infection, as the mouse and cell experiments represent models of severe illness, while the human cohorts consisted of individuals where SARS-CoV-2 infection was generally well tolerated and many subjects were entering the recovery phase. Studies have provided mechanistic evidence for metabolic dysregulation in COVID-19, notably through insulin resistance involving adiponectin/leptin and proinflammatory alterations, (44, 45) which fits with our observations, along with the decreased food intake observed during the acute phase of human disease, i.e. as evidenced by the infected mice in our studies (45).

A novel finding from our patient cohort is the separation of the COVID.POS patients into a sub-group with low/undetectable viral load on the day of saliva collection based on their saliva FTIR spectra. Saliva FTIR spectra of this COVID.POS^FU.NEG^ group displayed reduced saccharide (ribose) bands (1038 cm^-1^, 1074 cm^-1^) compared with COVID.POS^FU.POS^. These bands coincided with the ATR-FTIR bands for extracted SARS-CoV-2 RNA(46), in agreement with the PCR results; however, this difference in saccharide absorbance may also indicate recovery from the previously described, hyperglycemic state This group also showed reduced signal in the finger print region, proposed by Martinez-Cuazitl et al.(4) to represent immunoglobulins IgG, IgM and IgA. As these COVID.POS^FU.NEG^ patients are likely to have continued immunoglobulin expression/secretion (47), our results suggest that the Amide I/II and fingerprint regions more likely correlates with clearance of protein aggregates (β-sheet) and aliphatic amino acids as seen by significant decreases at 1688 cm^-1^ and 1373 cm^-1,^ respectively (Fig. 4b).

While the predictive model using saliva ATR-FTIR spectra showed high sensitivity in predicting COVID.NEG and COVID.POS^FU.POS^ cases, it was unable to differentiate COVID.POS^FU.NEG^ cases accurately. Nevertheless, when thinking how one may utilize this assay, as a possible point-of-care screening test, the high sensitivity makes it able to rule out infected individuals who are likely to transmit SARS-CoV-2, which is extremely important from an epidemiological and social perspective, particularly in the context of family groups when a child or one parent is infected, but other family members are not. This feature differentiates the FTIR saliva test from other point-of-care test with high specificity testing characteristics, making FTIR preferable especially in settings of large gatherings thereby potentially circumventing a super spreader event(s). One additional application for this assay is the ability to monitor physiological responses to COVID-19 which in turn may inform patient infection responses and predictions on prognosis and need for therapeutic interventions.

In contrast to previous fasting requirements of >8-hr prior to saliva collection (4), we took a pragmatic approach of only 20-30 minutes abstinence from food prior to testing. Our results support this time interval between sample collection and testing making a point-of-care rapid testing application more feasible. It is unlikely that this time interval can be shortened further as saliva is likely to be “contaminated” with food particles interfering with FTIR signals. We did notice, however, excessive precipitation while mixing saliva with ethanol, secondary to the initial high postprandial cephalic secretion. Adding a low-speed centrifugation of raw saliva prior to inactivation with ethanol circumvented this problem. Our simple, inactivation procedure with ethanol removes any possible biosafety concerns. All these features make future development for point-of-care application feasible. However, saliva collection and processing methods would require additional refinement (e.g. use of a capillary action sampling cartridge).

In conclusion, ATR-FTIR technology with saliva self-collection provides a simple, rapid and biosafe sample processing, which has high potential as a non-invasive, low-resource method for COVID-19 screening. The simplicity of the method means that only basic skills are required to conduct the test, which would satisfy the global need for rapid COVID-19 screening at diverse locations such as airports and public venues. Further evaluation may also establish utility for COVID-19 prognosis. As the method requires only generic laboratory equipment, ethanol, an ATR-FTIR with implemented predictive algorithm, and a power source, it offers promise as a global tool in the management of COVID-19 pandemic.

## Materials and methods

### SARS-CoV-2 virus

The SARS-CoV-2 isolate (hCoV-19/Australia/QLD02/2020) was kindly provided by Dr Alyssa Pyke (Queensland Health Forensic & Scientific Services, Queensland Department of Health, Brisbane, Australia). Virus stocks were prepared in Vero E6 cells as described (48) with all infectious SARS-CoV-2 work conducted in a dedicated suite in a biosafety level-3 (PC3) facility at the QIMR Berghofer MRI (Australian Department of Agriculture, Water and the Environment certification Q2326 and Office of the Gene Technology Regulator certification 3445). An aliquot of the viral stock was extracted and sequenced using illumina technology and uploaded to GISAID (https://www.gisaid.org/) under Accession ID (EPL_ISL_407896).

### *In vitro* cell model

Vero E6 (C1008, ECACC, Wiltshire, England; obtained via Sigma Aldrich) were maintained in RPMI 1640 (Thermo Fisher Scientific), supplemented with endotoxin free 10% heat-inactivated fetal bovine serum (FBS; Sigma-Aldrich), at 37°C and 5% CO_2_. Cells were checked for mycoplasma using MycoAlert Mycoplasma Detection Kit (Lonza, Basel, Switzerland). FBS was checked for endotoxin contamination before purchase as described (49).

Vero E6 cells (6×10^5^) were plated onto 6-well plates in 2ml RPMI + 10% FBS without phenol red (to limit background spectra). Following 24h of growth, media was removed and cells were subjected to control, mock-infection or infection regimes, each in triplicate. Control (untreated) cells were rinsed 2xPBS and placed in 3ml phenol-red RMPI + 2% FBS. Mock-infected cells were incubated with 500 μL of UV-inactivated SARS-CoV-2 stock for 30 min, while infected cells were incubated with 500 μL SARS-CoV-2 viral stock MOI 0.01 for 30 min. Mock-infected and infected cells were then rinsed 2xPBS and placed in 3ml phenol-red RPMI + 2% FBS. Aliquots of conditioned media were collected at 24h and 48 h post-infection. At each time point, 100 μL of conditioned media was mixed with 300 μL ice cold 100% ethanol for FTIR, while 200 μL conditioned media was mixed with 600 μL Trizol-LS for PCR. At the 48h time point, remaining supernatant was discarded and cells were harvested in 400 μL Trizol-LS for PCR.

### Nucleic acid extraction and RT–qPCR

RNA was purified from tissue culture supernatants and saliva (TPCH and QIMRB cohorts) using Direct-zol RNA microprep kits (Zymo Research) and cDNA was generated using iScript™ Reverse Transcription Supermix (BioRad). For qPCR, SsoAdvanced™ Universal SYBR® Green Supermix (Bio-Rad) was used with two previously published primer sets targeting different regions of SARS-CoV-2: 1) Forward (5’-CAATGCTGCAATCGTGCTAC-3’) and reverse (5’-GTTGCGACTACGTGATGAGG-3’) primers targeting the N-gene; 2) Forward (5’-ACCTTCCCAGGTAACAAACCA-3’) and reverse (5’-TTACCTTTCGGTCACACCCG-3’) primers targeting the 5’UTR. Cycling was carried out in a CFX384 Touch™ Real-Time PCR Detection System (Bio-Rad) under the following conditions: 95°C 30s; 95°C 10s, 60°C 30s (40x); melt curve 65°C-95°C. Viral copy number in experimental samples was estimated relative to a reference cDNA standard, using primer set 1. The reference cDNA was generated from a pool of SARS-CoV-2 infected VERO-E6 cell supernatant RNA and the viral copy number of reference cDNA was estimated relative to a plasmid containing the 5’UTR of SARS-CoV-2 (gift from Dongsheng Li, QIMR Berghofer MRI), using primer set 2. Plasmid copy number was determined using the URI Genomics and Sequencing Centre online calculator (http://cels.uri.edu/gsc/cndna.html). Saliva RNA quality was confirmed by amplification of housekeeping gene, β2-microglobulin, using forward (5’-ACTCTCTCTCTTTCTGGCCTGG-3’) and reverse (5’-CATTCTCTGCTGGATGACGTG-3’) primers.

### Mouse model

All mouse work was conducted in accordance with the “Australian code for the care and use of animals for scientific purposes” as defined by the National Health and Medical Research Council of Australia. Mouse work was approved by the QIMR Berghofer Medical Research Institute animal ethics committee (P3600, A2003-607). K18-hACE2+/- mice were purchased from Jackson laboratories and were maintained in-house as heterozygotes by backcrossing to C57BL6/J mice (17, 48). Mice were typed as described (48) using hACE2 Primers: Forward: 5’-CTT GGT GAT ATG TGG GGT AGA -3’; Reverse: 5’-CGC TTC ATC TCC CAC CAC TT -3’ (recommended by NIOBIOHN, Osaka, Japan).

Prior to oral lavage, 4-5 months old K18-hACE2+/- mice were placed in static micro isolator cages (Techniplast Static Micro-isolator # 1264) with gridfloor accessory, allowing feces to pass through for 1 hour without food or water, to avoid fecal contamination in oral cavity. Oral lavage was conducted with the mice under light anesthesia: 3% isoflurane (Piramal Enterprises Ltd., Andhra Pradesh, India) delivered using The Stinger, Rodent Anesthesia System (Advanced Anaesthesia Specialists/Darvall, Gladesville, NSW, Australia). With the mouse lying on its back, 25 μL of milliQ water was placed into the side of the mouth just behind the teeth of the lower mandible. The water was pipetted up and down 4 times to wash the mouth without injury or abrasion of gums or lips. The lavage was recovered, and 15 μL was added to 45 μL of 100% ethanol to obtain 75% v/v ethanol, then stored at -80ºC.

Mice were infected intrapulmonary via the nasal route with 5×10^4^ CCID_50_ SARS-CoV-2 in 50 μL medium while under light anesthesia. Saliva samples were collected before infection and on the indicated days after infection. Mice body weights were measured each day. Mice were euthanized using CO_2_ on day 4 post infection and lung titers determined by CCID_50_ assay of serial dilution of supernatants from homogenized lung tissues (48).

### Mouse Lavage Proteomics

Protein was extracted from ethanol-containing lavage samples by centrifugation at 16,000x g for 25 minutes, at 4°C. Supernatant was discarded, and protein pellet washed twice with 50 mM triethylammonium bicarbonate (TEAB, Sigma-Aldrich) buffer. Proteins were resuspended in 50 mM TEAB and underwent protein estimation by BCA assay, per manufacturer’s instruction (Thermo Fisher Scientific). Outliers were defined as samples with a protein abundance 3 standard deviations above the mean for that condition, and were excluded from further processing. The resulting day 0 lavage protein samples were pooled due to their low abundance resulting in a single proteomics sample. Day 4 lavage samples contained adequate protein abundance to continue as individual replicates, n = 4 for SARS-CoV-2^UV-I^ and n = 8 for SARS-CoV-2^POS^ conditions. The BRAVO AssayMap platform (Agilent Technologies) was used for in-solution digest and C18 desalting procedures. 1% sodium deoxycholate was added to each sample for increased protein solubility. A standard automated trypsin digest method was followed using 5 mM dithiothreitol and 20 mM 2-iodoacetamide. Samples were diluted 1:10 with 50 mM TEAB and porcine trypsin (Promega) added (final 1:30 trypsin to sample protein ratio). Digests incubated overnight at 37°C and acidified using trifluoroacetic acid (TFA) to a final concentration of 0.5%. Sodium deoxycholate was pelleted by centrifugation for 30 min, at 5,000x g, room temperature. Peptides were subsequently desalted by AssayMAP C18 cartridge, following manufacture’s instruction. Eluted peptide was dried and resuspended in 0.5% TFA. Peptides were resolved on a Thermo U3000 nanoHPLC system and analysed on a Thermo Q Exactive Plus Orbitrap mass spectrometer. The HPLC setup used a C18 trap column and a 50 cm EasySpray C-18 analytical column (Thermo Fisher, catalogue: 160454, ES803A). Mobile phases were A: 0.1% formic acid, and B: 80% acetonitrile with 0.1% formic acid. The loading pump ran on 3% B at 10 μL per minute. 1 μg peptide were loaded in 3% B. The nano-capillary pump ran at 250 nL per minute, starting at 3% B. The multi-step gradient was 3% to 6% B over 1 minute, 6% to 30% B over the following 60 minutes, 30% to 50% B over the following 12 minutes, then 50% to 95% B over 1 minute. After maintaining 95% B for 12 minutes, the system was re-equilibrated to 3% B. The mass spectrometer ran an EasySpray source in positive ion DDA mode, using settings typical for high complexity peptide analyses. Mass lock was set to “Best”. Full MS scans from 350 m/z to 1400 m/z were acquired at 70k resolution, with an AGC target of 3E6 and 100 ms maximum injection time. MS2 fragmentation was carried out on the Top 10 precursors, excluding 1+ and > 7+ charged precursors. The dynamic exclusion window was 30 seconds. Precursor isolation width was 1.4 m/z and NCE was 27. MS2 resolution was 17,500, with an AGC target of 5E5 and a maximum injection time of 50 ms. Protein identification was completed by MaxQuant using Swiss-Prot mouse proteome (version 2021_04) and default parameters. Label-free quantitation intensities were analysed by the LFQ-Analyst pipeline to determine differentially abundant proteins based on p-values < 0.1 (Benjamini Hochberg adjusted p-value). Intensities were Z-score normalized and expressed as a heat map.

### Cohort study

The project was approved by Human Research Ethics Committees of QIMR Berghofer Medical Research Institute (QIMRB, P3675), New South Wales Health Pathology (NSWHP-RPAH 2020/ETH02630) and The Prince Charles Hospital (TPCH, ID 63003). All participants provided written informed consent.

The cohort originated from three sites and included i) asymptomatic healthy volunteers (QIMRB and TPCH) not in contact with COVID-19 cases for past 14 days (COVID.NEG); ii) COVID-19 positive (COVID.POS) hospitalised patients at TPCH, and iii) COVID.POS individuals in hotel quarantine in New South Wales, Sydney, Australia. For cohorts ii) and iii) saliva sampling was performed within 14 days after the initial PCR diagnostic test.

Fasting prior to saliva collection was considered but dropped to align with the real-world screening scenario. Participants were requested to rinse mouth with water and refrain from eating and drinking for 20 minutes prior to collecting 1.2 to 3 ml saliva as sublingual drool into a clean receptacle.

Samples from cohorts i) and ii) were stored on ice and processed within 30 minutes. After brief vortex, an aliquot of raw saliva was transferred to a 1.5 ml Eppendorf tube and centrifuged for 10 minutes at 500x g at 4 C to remove particulates. Clarified saliva was transferred to a cryotube containing ethanol to obtain 75% v/v ethanol and incubated at room temperature for 30 minutes.

Inactivated saliva samples were stored at -80°C. For the TPCH COVID.POS samples, another tube was prepared to 75% v/v Trizol for RT-PCR using the protocol described above.

Samples from cohort iii) were initially transported to the laboratory at room temperature. Aliquots of raw saliva were frozen at -80°C, subsequently thawed on ice, inactivated with 75% v/v ethanol and shipped on dry-ice to QIMR Berghofer for FTIR analysis. A nasal pharyngeal swab was collected on the same day as saliva, and was analysed by RT-PCR using TaqPath COVID-19 Combo Kit (Thermo Fisher Scientific) according to manufacturer instructions.

For COVID.POS individuals, a subset tested PCR negative and were classified as COVID.POS^FU.NEG^ (Table S1).

### ATR-FTIR spectra acquisition and processing

Samples in 75% ethanol were thawed on ice and homogenised by high speed vortexing. An aliquot of 2 μL was applied to the crystal of an ATR-FTIR instrument (Agilent Cary 630). and allowed to air dry (∼30 sec) before spectral acquisition occurred over the wavenumber range, 4000-650 cm^-1^. Background was collected without sample, i.e. ambient room air at 21□ between each measurement following cleaning of the crystal with 80% ethanol. Settings included 64 scans (Sample/Background) with a resolution of 8 cm^-1^. All spectra were baseline adjusted with baseline estimated using regions 2031-1865 cm^-1^ and 3971-3799 cm^-1^. Spectra were then normalised by adjusting to area under the curve (AUC) as 1.

### Statistical analysis

Euclidean distance was calculated for each pairwise comparison of normalised spectra to determine intra- and inter-sample variability. Each comparison was grouped into a “intra-sample” (spectra from same biological replicate, 1,970 comparisons) or “inter-sample” (spectra from different biological replicate, 89,253 comparisons) category and represented as a violin plot.

Clustering of the samples was explored using discriminant analysis to create a canonical plot to display clustering of clinical groups. LogWorth statistic was applied to identify spectral regions that significantly deviate between two sample groups. The false discovery rate p-value cut-off for each comparison was chosen in a data-dependent manner accounting for the differences from baseline.

A predictive model was developed using Partial Least Squares Discriminant analysis (PLS-DA) to predict clinical group based on the spectra. A six factors solution was chosen to account for at least 70% of the variation in the spectrum. A variable importance plot (VIP) was generated to indicate what areas of the spectrum most contributed to the predictive model. The fit of the model was evaluated using Leave-One-Out Cross-validation (LOO-CV), generating a receiver operating characteristic (ROC) curve and a confusion matrix giving the cross validated sensitivity and specificity. The cutoff for predicting positive or negative were chosen to maximize Youden’s Index (ie. the sum of sensitivity and specificity).

## Data Availability

Raw FTIR spectra and associated clinical data are available at DOI: 10.5281/zenodo.5703689,
Proteomics data are available via ProteomeXchange with identifier PXD030012.

https://zenodo.org/record/5703689#.YZLxWGBBxaQ

## Data availability

Raw FTIR spectra and associated clinical data are available at DOI: 10.5281/zenodo.5703689, https://zenodo.org/record/5703689#.YZLxWGBBxaQ

Proteomics data are available via ProteomeXchange with identifier PXD030012.

## Acknowledgements

We thank all participants for donating their time and saliva samples to this research. From QIMR Berghofer Medical Research Institute we thank Dr Itaru Anraku for managing the PC3 (BSL3) facility and animal house staff for mouse breeding and agistment. We thank Dr Alyssa Pyke and Mr Fredrick Moore (Queensland Health, Brisbane) for providing the SARS-CoV-2 isolates. We thank Clive Berghofer and the Brazil Family Foundation (and many others) for their generous philanthropic donations to support SARS-CoV-2 research at QIMR Berghofer. We thank Peter Vardon, research nurse for coordinating the biological sample collection at The Prince Charles Hospital. A.S. holds an Investigator grant from the National Health and Medical Research Council (NHMRC) of Australia (APP1173880).

## Funding

This project was supported by Agilent Technologies Applications & Core Technology - University Research Grant (MMH), QIMR Berghofer Medical Research Institute COVID Research (AS, DR) and The Prince Charles Hospital Research Foundation (DR).

## Conflict of interest statement

AH, DB, FZ are employees of Agilent Technologies. All other authors declare that they have no competing interests.

## Supporting information

S1 Fig. Culture supernatant ATR-FTIR spectra and subtractive analysis of Vero cell SARS-CoV-2 infection model.

S2 Fig. Full ATR-FTIR spectra and subtraction analysis of mouse oral lavage.

S3 Fig. Significant features of in vivo infection mouse model using LogWorth FDR analysis.

S4 Fig. Acceptable technical variance between replicates using pairwise Euclidean distancing analysis of human samples.

S5 Fig. Human subject ATR-FTIR close-up of averaged spectra per three groups.

S1 Table. Summary of cohorts.

## AUTHOR CONTRIBUTIONS

Conceptualization: MMH, RSR, DB, AS

Data curation: GH, HR

Formal analysis: GH

Funding acquisition: DWR, AS, MMH

Investigation: STK, HR, RSR, KY, TTL

Methodology: HR, RSR, AH, FZ, DJR, AS, MMH

Project administration: RSR, DWR, MMH

Resources: SVH, RC, DWR

Supervision: AS, MMH

Visualization: HR

Writing – original draft: MMH, STK, HR

Writing – review & editing: GH, RSR, DJR, SVH, DWR, AH, DB

## Notes

### Author Declarations

Human research ethics committee of QIMR Berghofer Medical Research Institute gave ethical approval for this work.

