## Supplementary for "Infrared spectroscopy enables rapid, robust, portable COVID-19 saliva screening based on pathophysiological response to SARS-CoV-2"

**Contents**

S1 Fig. Culture supernatant ATR-FTIR spectra and subtractive analysis of Vero cell SARS-CoV-2 infection model.
S2 Fig. Full ATR-FTIR spectra and subtraction analysis of mouse oral lavage.
S3 Fig. Significant features of in vivo infection mouse model using LogWorth FDR analysis.
S4 Fig. Acceptable technical variance between replicates using pairwise Euclidean distancing analysis of human samples.
S5 Fig. Human subject ATR-FTIR close-up of averaged spectra per three groups.
S1 Table. Summary of cohorts.

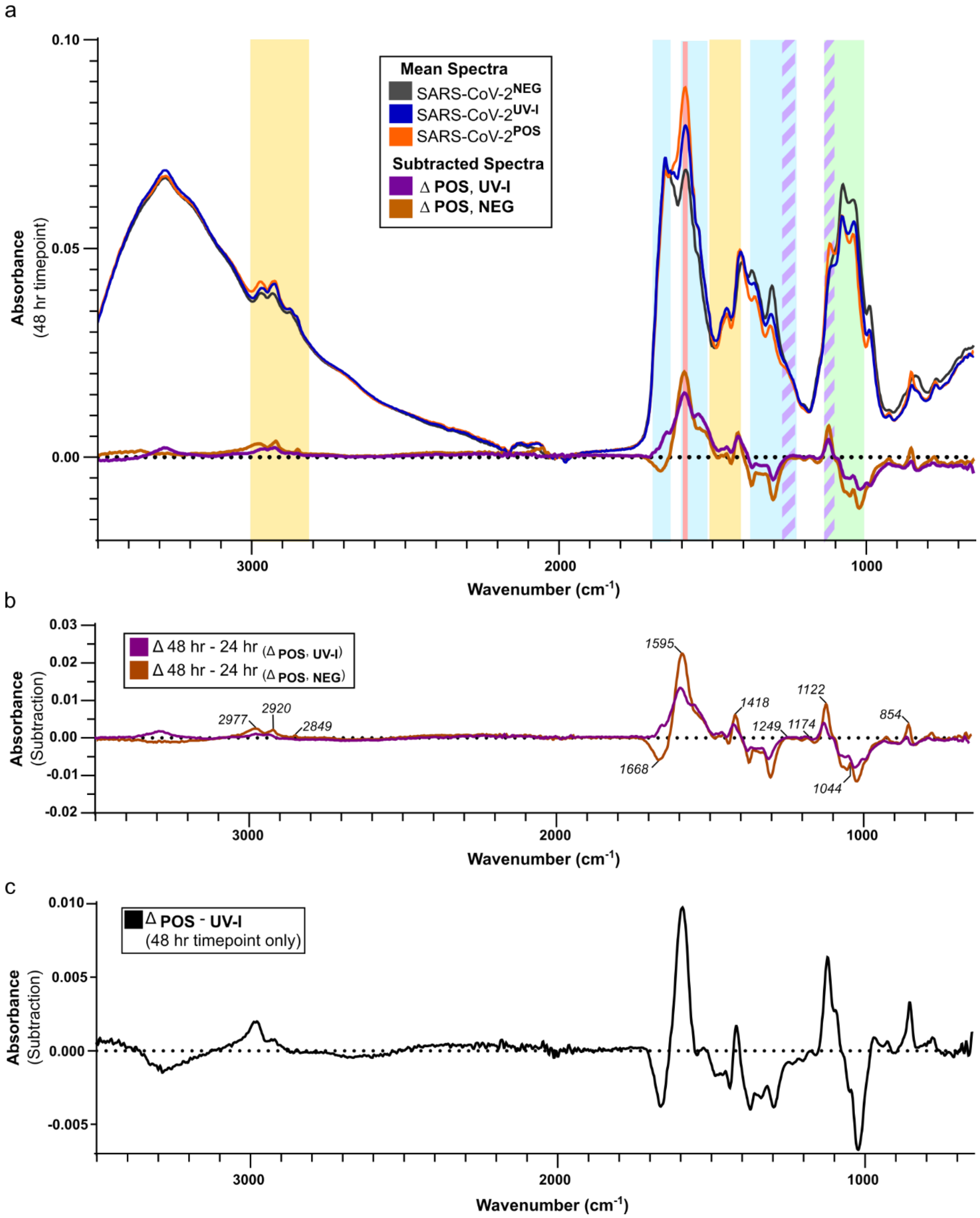


**S1 Fig. Culture supernatant ATR-FTIR spectra and subtractive analysis of Vero cell SARS-CoV-2 infection model.** Refer to Fig 1 for experimental details. **a.** Spectra of all three groups at 48 hr time point (grey, blue, orange) with subtracted spectra (Δspectra) along the X-axis, SARS-CoV-2 spectra subtracted from media control (dark orange), SARS-CoV-2 spectra subtracted from inactivated control (purple). Bands signified by colours: Aliphatic (yellow), Amide I/II/III (cyan), severity (red), Saccharide (green), phosphodiester asymmetric stretching ($\nu_{as}PO_2^-$) and symmetric stretching ($\nu_sPO_2^-$) (purple stripes). **b.** Isolated subtraction spectra (48 hr – 24 hr) of ΔSARS-CoV-2 from media control (dark orange) and inactivated control (purple). **c)** The unique spectral changes of active infection at 48 hr: spectra subtraction of SARS-CoV-2 from Inactivated SARS-CoV-2 at 48 hr.

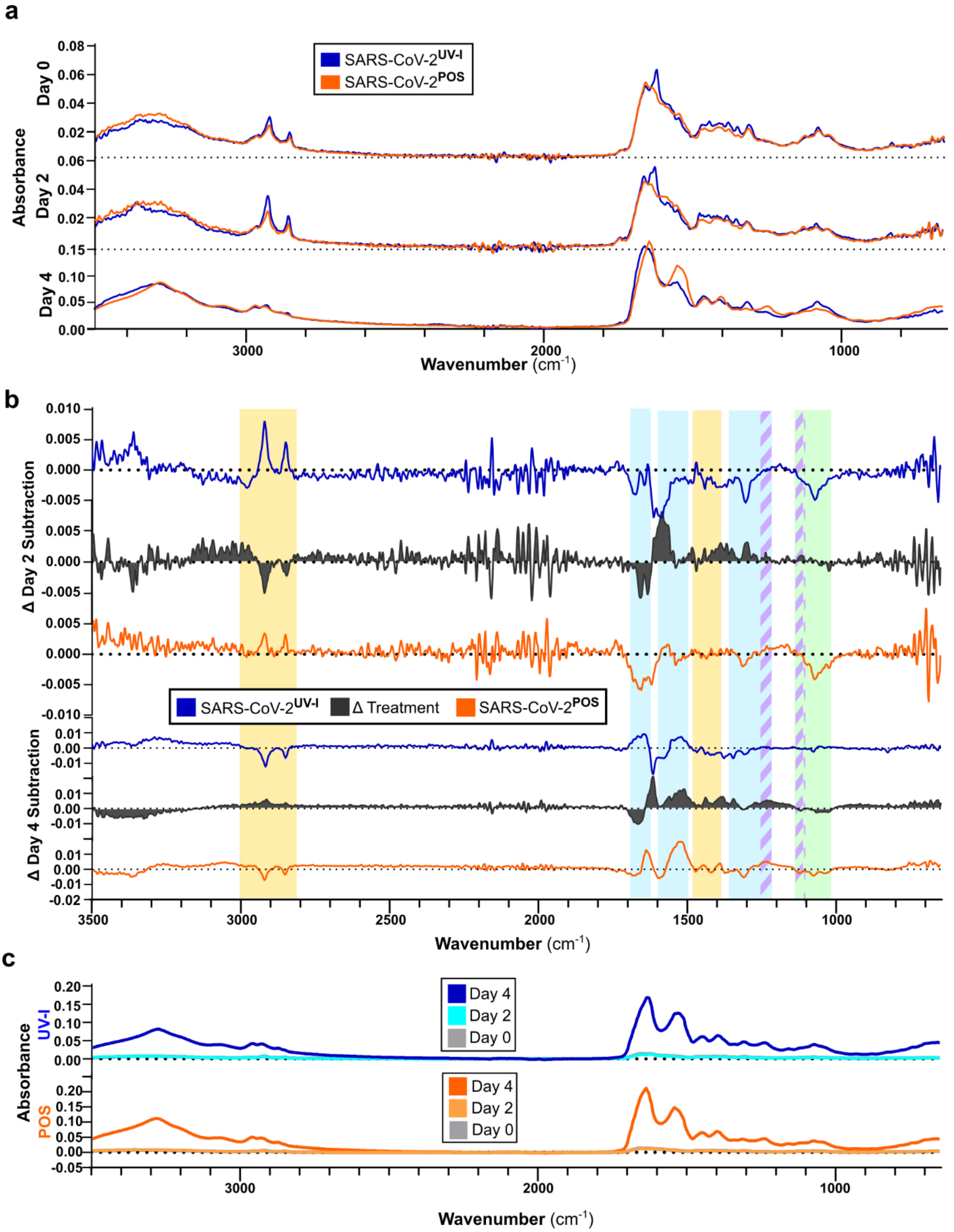


**S2 Fig. Full ATR-FTIR spectra and subtraction analysis of mouse oral lavage. a)** ATR-FTIR spectra (3500-650 cm$^{-1}$) of active SARS-CoV-2 (orange) and Inactivated SARS-CoV-2 (blue) groups per day 0 (baseline), day 2, and day 4. **b)** Spectra subtraction at days 2 and 4 from day 0 per each group. The 'Δ Treatment' (grey) condition is the difference between SARS-CoV-2 and Inactivated SARS-CoV-2 time-point subtractions as respectively illustrated. Bands signified by colours: Aliphatic (yellow), Amide I/II/III (cyan), Saccharide (green), phosphodiester asymmetric stretching ($\nu_{as}PO_2^-$) and symmetric stretching ($\nu_sPO_2^-$) (purple stripes). **c)** Un-normalized spectra showing overlapping spectra for days 0, 2, and 4, per each respective group: Inactivated SARS-CoV-2 (top), SARS-CoV-2 (bottom).

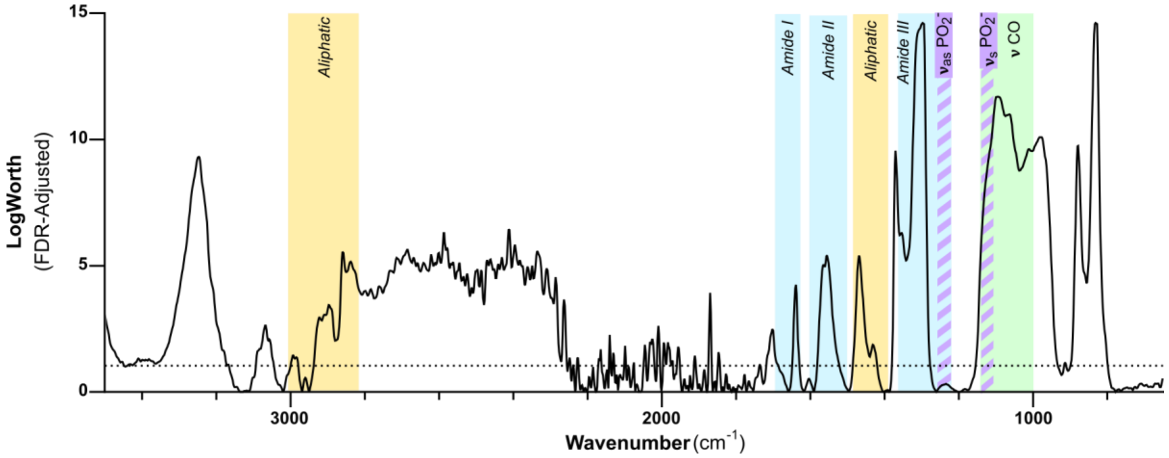


**S3 Fig. Significant features of in vivo infection mouse model using LogWorth FDR analysis.** All bands of interest demonstrated significant changes by day 4; dotted line indicating FDR=1.3 (P<0.05). Features of aliphatic signals, protein, nucleic acid (RNA), and saccharides were of all found to present significant alterations by SARS-CoV-2 infection.

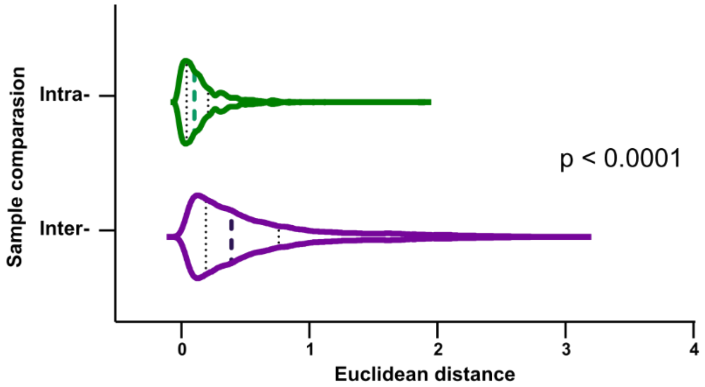


| | Inter | Intra |
|---|---|---|
| Number of values | 89,253 | 1,970 |
| Minimum | 0.0271 | 0.009915 |
| 25% Percentile | 0.2233 | 0.07259 |
| Median | 0.4231 | 0.1333 |
| 75% Percentile | 0.7929 | 0.2423 |
| Maximum | 3.126 | 1.92 |
| Range | 3.099 | 1.91 |
| Mean | **0.6089** | **0.1925** |
| Std. Deviation | 0.544 | 0.1941 |
| Std. Error of Mean | 0.001821 | 0.004373 |

**S4 Fig. Acceptable technical variance between replicates using pairwise Euclidean distancing analysis of human samples.** Analysis comparing normalized FTIR spectra variance of technical replicates (Intra) to that of biological (Inter). A statistical difference of $p < 0.0001$ was observed when comparing distributions of the two groups.

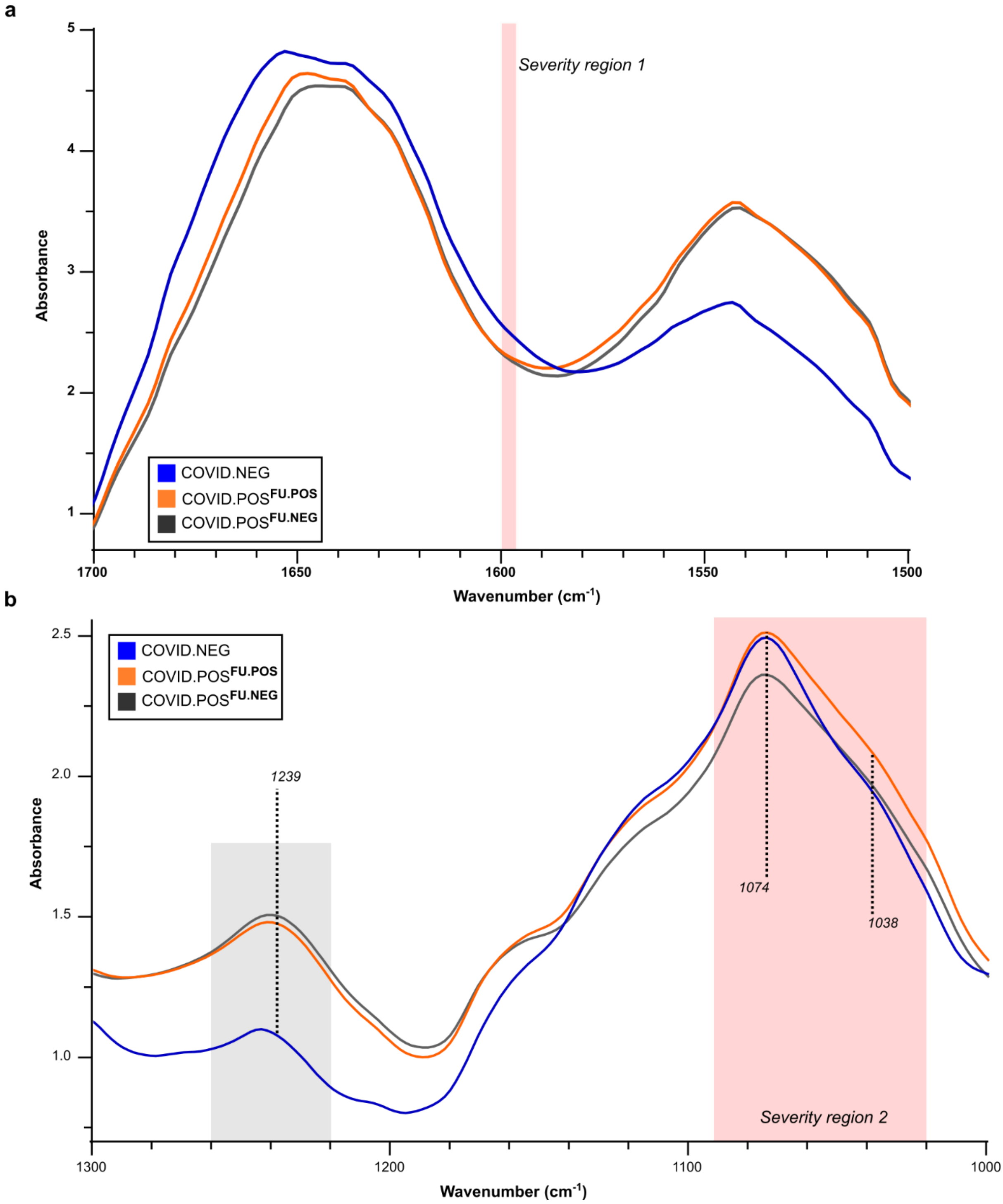


**S5 Fig. Human subject ATR-FTIR close-up of averaged spectra per three groups.** a) Magnified view of the averaged human spectra of the three groups along the range 1700-1500 $cm^{-1}$. Groups are identified as Post infection (grey), Healthy control (blue), and SARS-CoV-2 positive (orange). b) The Amide III and saccharide bands magnified along the range, 1300-1000 $cm^{-1}$. Wavenumbers labelled as 1239, 1074, and 1038 [1], have been shown to represent SARS-CoV-2 RNA, while 1260-1220 $cm^{-1}$ (grey) also represents protein amyloid aggregates.[2]

**S1 Table. Summary of cohorts**

| | i) QIMRB/TPCH | ii) TPCH | iii) NSW HP |
|---|---|---|---|
| Healthy | 42/2 | 0 | 0 |
| COVID.POS | | | |
| Saliva PCR positive | 0 | 6 | 40 |
| Saliva PCR negative | 0 | 4 | 10 |
| **Total (n)** | **44** | **10** | **50** |

TPCH, The Prince Charles Hospital; QIMRB, QIMR Berghofer Medical Research Institute; NSWHP, New South Wales Health Pathology
